# Audiometric hearing thresholds and operational cardiovascular-kidney-metabolic staging in U.S. adults aged 70-79 years: a cross-sectional NHANES analysis

**DOI:** 10.64898/2026.09.11.26362649

**Authors:** Dinh Minh Le, Son Truc Tran, Van Chien Nguyen

## Abstract

**Objective:** To examine whether bilateral four-frequency pure-tone average (PTA4) increased across operational cardiovascular-kidney-metabolic (CKM) stages and assess associations with individual CKM components.

**Design:** Cross-sectional National Health and Nutrition Examination Survey (NHANES) 2017-March 2020 pre-pandemic analysis. Survey-weighted models adjusted for demographics, education, smoking, and noise exposure.

**Study sample:** Adults aged 70-79 years (n = 994); stage analysis n = 285; waist model n = 778.

**Results:** Covariate-adjusted mean bilateral PTA4 thresholds did not increase consistently across stages. No planned Stage 2-4 comparison met the Holm criterion; Stage 4 minus Stage 3 was 4.45 dB HL (95% CI, − 0.61 to 9.51; Holm p = 0.246). Stage 2-4-only heterogeneity was not supported (p = 0.197). The five-stage omnibus was significant (p < 0.001), but inference was limited by sparse lower-stage support, including one Stage 0 participant. Waist circumference alone met the nine-test Holm criterion: +1.57 dB HL per 10 cm (95% CI, 0.74 to 2.39; Holm p = 0.00547).

**Conclusions:** Greater waist circumference was associated with higher PTA4, but this cross-sectional association does not establish a hearing benefit from reducing waist circumference. These data do not establish operational CKM stage as a graded indicator of PTA4 in adults aged 70-79 years.

## 1 Introduction

Hearing loss is common among older U.S. adults (Goman and Lin 2016), and audiometric thresholds are higher in older age groups (Humes 2023). Cardiovascular, kidney, and metabolic health may also be relevant to hearing in later life. Interpreting these relationships requires attention to how hearing is assessed. Pure-tone audiometry measures hearing sensitivity at specified frequencies, whereas self-reported hearing difficulty reflects perceived hearing function (de Gruy et al. 2023). Evidence concerning one outcome therefore may not directly answer questions about the other.

Observational syntheses have reported associations of hearing loss with diabetes (Nisar et al. 2026), hypertension (Jin et al. 2025), chronic kidney disease (Li et al. 2026), and adiposity (Yang et al. 2020). Studies examining broader cardiovascular health have used different exposures and hearing outcomes. In the HUNT study, several cardiovascular risk factors, including waist circumference, were associated with poorer hearing sensitivity, although their combined contribution was small and of uncertain clinical relevance (Engdahl et al. 2015). A Finnish population-based study did not detect a statistically significant association between cardiovascular disease and hearing impairment (Lohi et al. 2015), and its 13-year follow-up did not detect a statistically significant association between cardiovascular disease and subsequent hearing-threshold deterioration (Lohi et al. 2022). More recently, cardiovascular-kidney-metabolic (CKM) Stage 4 was associated with higher odds of incident self-reported hearing impairment than Stage 0 among Chinese adults aged 45 years or older (Zhang et al. 2026). Improvement in Life’s Essential 8 was also associated with lower odds of hearing loss under four-frequency and high-frequency better-ear definitions (Yévenes-Briones et al. 2026). These studies extend the evidence beyond individual conditions, but their exposures and outcomes address different aspects of the relationship between systemic health and hearing.

The CKM framework groups cardiovascular, kidney, and metabolic abnormalities into an ordered stage hierarchy (Ndumele et al. 2026). Whether this ordering corresponds to progressively higher continuous audiometric thresholds remains uncertain. The longitudinal CKM study assessed incident self-reported hearing impairment (Zhang et al. 2026), while the Life’s Essential 8 study examined change in cardiovascular health rather than CKM stage (Yévenes-Briones et al. 2026). Neither directly addressed whether mean continuous audiometric thresholds increased across CKM stages. Variation within stage groups provides a further reason to examine individual components. Participants can reach the same stage through different combinations of abnormalities, and their component measurements may differ despite a shared stage assignment (Ndumele et al. 2026). Stage comparisons can assess whether mean hearing thresholds follow the ordered hierarchy, while component analyses can describe associations with individual measurements. These complementary questions allow the relationship with hearing to be examined without assuming that an association with one component explains the pattern across stages or provides information incremental to stage.

We used National Health and Nutrition Examination Survey (NHANES) 2017-March 2020 pre-pandemic data to study U.S. civilian noninstitutionalized adults aged 70-79 years (Stierman et al. 2021). NHANES includes standardized audiometry (National Center for Health Statistics 2022) alongside interview, examination, and laboratory measures, allowing stage-level and component-level questions to be examined using the same hearing outcome. We assigned operational CKM stages from available measures. Our primary objective was to examine whether covariate-adjusted mean bilateral four-frequency puretone average (PTA4) increased across CKM stages. Secondary analyses examined associations between individual CKM components and PTA4. Exploratory analyses assessed whether selected component-PTA4 associations differed across stages.

## 2 Methods

### 2.1 Study design and target population

We conducted a cross-sectional analysis of public-use NHANES 2017-March 2020 pre-pandemic data. The target population was U.S. civilian noninstitutionalized adults aged 70-79 years. Adults aged 70 years and older were eligible for audiometry in this NHANES cycle (National Center for Health Statistics 2022). The upper age bound of 79 years matched the age range in which the Predicting Risk of Cardiovascular Disease Events (PREVENT) equations were developed and validated (Khan et al. 2024). Survey design objects were defined before applying age restrictions or model-specific eligibility criteria so that the released strata and primary sampling units were retained, consistent with NHANES analytic guidance (Akinbami et al. 2022).

The target-age domain included 994 participants. For the primary CKM stage analysis, participants additionally required a positive NHANES fasting-subsample survey weight; a resolved operational CKM stage; bilateral PTA4; and complete data for the seven common adjustment covariates. Component models used either mobile examination center (MEC) weights or fasting-subsample weights according to the variables included in each analysis. No a priori sample-size calculation was performed. Eligible participants were included according to each model’s survey-weight requirement and availability of exposure, outcome, and covariate data.

### 2.2 Audiometric outcome

The primary hearing outcome was bilateral PTA4, derived from air-conduction thresholds. Bilateral PTA4 was prespecified to summarize hearing-threshold sensitivity across both ears and four frequencies. For each ear, PTA4 was calculated as the mean hearing threshold at 0.5, 1, 2, and 4 kHz; bilateral PTA4 was then calculated as the mean of the right- and left-ear PTA4 values. Valid thresholds at all four frequencies were required for each ear, and bilateral PTA4 required both ears.

NHANES codes 666 as no response at the audiometric output limit and 888 as “could not obtain”; repeat and insert/crossover-retest thresholds are also released (National Center for Health Statistics 2022). Final code 666 was assigned 126 decibels hearing level (dB HL) as an analytic no-response convention; code 888 was treated as missing. Routine 1-kHz pairs were accepted when numeric values differed by ≤ 10 dB (first value retained) or both were 666; other pairs were unresolved. Insert/crossover retests replaced corresponding standard thresholds. Full audiometric rules are provided in Supplement Section S5.

### 2.3 Operational CKM stage

We assessed CKM criteria from Stage 4 down to Stage 0. Each stage-defining criterion was recorded as present, absent, or unresolved when the available data were insufficient (Table 1). Participants were assigned to Stage 0 only when the available data ruled out criteria for Stages 1-4 and showed the required normal findings for Stage 0. Stage-defining variables were not multiply imputed; unresolved evidence for a higher stage could prevent assignment to a lower stage. Full definitions, source variables, and missing-data rules are provided in Supplement Tables S2-S3.

**Table 1.** Operational CKM stage hierarchy used in this study.

| Stage | Operational rule |
| --- | --- |
| 0 | Resolved absence of Stages 1-4 together with all required normal Stage 0 findings; not a residual category. |
| 1 | Excess adiposity or prediabetes, with higher-stage criteria resolved as absent. |
| 2 | Hypertension, hypertriglyceridemia, metabolic syndrome, operational diabetes, or moderate or higher kidney risk. |
| 3 | Very-high kidney risk or PREVENT 10-year total cardiovascular disease risk $\geq 20\%$, without observed clinical cardiovascular disease. |
| 4 | Observed clinical cardiovascular disease together with definite operational CKM context; exact distinction between prediabetes and operational diabetes was not required to establish context. |

*Note.* Stage assignment was hierarchical ( $4 > 3 > 2 > 1 > 0$ ) and preserved unresolved higher-stage evidence. Stage 0 was not a residual category. CKM = cardiovascular-kidney-metabolic; PREVENT = Predicting Risk of Cardiovascular Disease Events.

Stage 1 included excess adiposity or prediabetes. Excess adiposity was body mass index (BMI) ≥ 25 kg/m^2^ or waist circumference ≥ 102 cm in men and ≥ 88 cm in women; for non-Hispanic Asian participants, thresholds were BMI ≥ 23 kg/m^2^ or waist circumference ≥ 90 cm in men and ≥ 80 cm in women (Ndumele et al. 2026). For the public-use “Other Race-Including Multi-Racial” category, concordant results under both threshold systems were classified and discordant results were unresolved. Bounding checks using either threshold system for the whole category did not change final stage assignments. Operational diabetes was defined by clinician-diagnosed diabetes, hemoglobin A1c (HbA1c) ≥ 6.5%, or fasting plasma glucose ≥ 126 mg/dL (Ndumele et al. 2026). Fasting glucose was used only among participants with a positive fasting-subsample weight. Prediabetes was evaluated only after operational diabetes had been excluded and was defined by HbA1c of 5.7%-6.4% or fasting plasma glucose of 100 to <126 mg/dL (Ndumele et al. 2026). The same positive fasting-subsample weight requirement was used for fasting triglycerides.

Stage 2 included hypertension, hypertriglyceridemia, metabolic syndrome, operational diabetes, or moderate or higher kidney risk (Ndumele et al. 2026). Hypertension was defined by mean systolic blood pressure (SBP) ≥ 130 mmHg or mean diastolic blood pressure (DBP) ≥ 80 mmHg, calculated from at least two valid measurements, or by current prescribed anti-hypertensive treatment. Hypertriglyceridemia required fasting triglycerides ≥ 150 mg/dL with a positive fasting-subsample weight. Metabolic syndrome required at least three of the five operational criteria to be definitely positive. Criterion-level definitions and missing-data rules are provided in Supplement Table S3.

Stage 3 required either very-high kidney risk or PREVENT 10-year total cardiovascular disease risk ≥ 20% in the absence of observed clinical cardiovascular disease (Ndumele et al. 2026). The PREVENT equations were developed and validated in adults aged 30-79 years without known cardiovascular disease and include diabetes as a binary predictor (Khan et al. 2024). For the study implementation, that binary diabetes input was based on clinician-diagnosed diabetes history, whereas the broader laboratory-inclusive diabetes definition was used elsewhere in CKM staging. Using diabetes history rather than the laboratory-inclusive definition could change which participants were assigned to Stage 3. PREVENT risk was not calculated when a required input was missing or fell outside the accepted implementation range.

The clinical CKM framework defines Stage 4 by clinical cardiovascular disease in the setting of CKM syndrome (Ndumele et al. 2026). Operational Stage 4 required observed clinical cardiovascular disease plus confirmed CKM context from excess adiposity, dysglycemia, hypertension, hypertriglyceridemia, metabolic syndrome, or moderate or higher kidney risk. Observed clinical cardiovascular disease comprised self-reported clinician diagnoses of heart failure, coronary heart disease, angina, myocardial infarction, or stroke. Peripheral artery disease and atrial fibrillation were not comprehensively captured, and direct subclinical cardiovascular measures were unavailable.

Estimated glomerular filtration rate (eGFR) was calculated using the 2021 Chronic Kidney Disease Epidemiology Collaboration creatinine equation without race (Inker et al. 2021). eGFR and urine albumin-to-creatinine ratio (UACR) were then mapped to Kidney Disease: Improving Global Outcomes (KDIGO) kidney-risk regions (Kidney Disease: Improving Global Outcomes (KDIGO) CKD Work Group 2024). Because NHANES provides these measurements from a single examination, an abnormal eGFR or UACR value could not establish the chronicity required for a clinical diagnosis of chronic kidney disease (Kidney Disease: Improving Global Outcomes (KDIGO) CKD Work Group 2024). The operational stage hierarchy is summarized in Table 1.

### 2.4 Analytic framework and CKM-related component analyses

H1 was the primary stage analysis and asked whether higher operational CKM stages were associated with higher bilateral PTA4; stage was modeled categorically. Its three planned pairwise contrasts compared adjusted mean bilateral PTA4 between Stage 3 and Stage 2, Stage 4 and Stage 2, and Stage 4 and Stage 3. H2 was a secondary analysis of associations between individual CKM components and PTA4. H3 examined whether selected component-PTA4 associations differed across stages; the Stage 2-4 interaction analyses were post-hoc and exploratory. H4 examined selected within-stage phenotypes and was developed after hearing-outcome review; it was post-hoc and hypothesis-generating. The timing of the staging correction and its implications for H1 are described in Supplement Section S1.

H2 examined nine CKM-related measures in separate survey-weighted models: BMI, waist circumference, SBP, DBP, HbA1c, eGFR, UACR, fasting triglycerides, and high-density lipoprotein cholesterol (HDL-C). Associations were reported per 5 kg/m^2^ higher BMI, 10 cm greater waist circumference, 10 mmHg higher SBP or DBP, 1 percentage point higher HbA1c, 10 mL/min/1.73 m^2^ lower eGFR, and 10 mg/dL higher HDL-C. UACR and triglycerides were modeled on the log2 scale and reported per doubling. Eight models used examination weights and triglycerides used fasting-subsample weights. Components were not mutually adjusted, ranked against one another, or tested for incremental information beyond CKM stage.

H3 evaluated whether associations of waist circumference, SBP, HbA1c, eGFR, UACR, and HDL-C with PTA4 differed across CKM stages. Each model included CKM stage, the continuous component, a stage-by-component interaction, and the seven common adjustment covariates. The joint interaction test evaluated the null hypothesis that the component slopes were equal across the included stages. The original five-stage interaction models were structurally non-estimable because the single Stage 0 participant could not support a within-stage continuous slope. The same six interaction tests were therefore evaluated post-hoc within Stages 2-4.

### 2.5 Post-hoc within-stage phenotype analyses

After review of the hearing-outcome results, H4 was developed as an additional exploratory analysis to examine associations between selected within-stage phenotypes and PTA4. H4 was not part of the original H1-H3 analytic hierarchy and was treated as post-hoc and hypothesis-generating. Two separate Holm multiplicity families tested (1) whether stage-specific coefficients for one characteristic were jointly zero across included stages and (2) whether the coefficients for a set of characteristics were jointly zero within a stage.

The parameter-level family paired hypertension duration with current SBP (Stages 2-4), diabetes duration with HbA1c (Stages 3-4), and waist circumference with derived 10-year weight change (Stages 2-4). Each characteristic had a separate coefficient for each included stage. One joint test was performed for each characteristic, giving six tests across the three paired models in one Holm multiplicity family. These tests asked whether the coefficients for a characteristic were jointly zero while its paired characteristic remained in the model. Between-stage differences in component slopes were examined separately in the H3 interaction models.

The multidimensional family tested characteristic blocks within stages. Stage 3 and 4 hypertension models included hypertension duration, current SBP, medication-class composition, and current-treatment duration from prescription records (National Center for Health Statistics 2021). Stage 2-4 adiposity models included waist circumference, derived 10-year weight change, and self-reported weight at age 25. The two hypertension and three adiposity block tests formed a second Holm multiplicity family.

Medication-class composition (one versus two or more anti-hypertensive classes) was an observed phenotype marker, not a measure of treatment intensity, hypertension severity, or resistant hypertension. H4 used fasting-subsample weights and the common adjustment set. Diagnosis-age handling is detailed in Supplement Section S4.5, model support in Supplement Table S18, representative-profile methods in Supplement Section S15, and secondary kidney/lipid analyses in Supplement Section S17.

### 2.6 Covariates and statistical analysis

The common adjustment set was age, sex, race/ethnicity, education, smoking status, occupational noise exposure, and nonoccupational loud-noise exposure and was retained across H1-H4. Education entered as five NHANES educational-attainment categories; age entered continuously and the other covariates categorically. Age, sex, and smoking also contribute to PREVENT-based Stage 3 assignment, so adjusted stage comparisons condition on some information used in staging. Survey-weighted linear regression modeled bilateral PTA4 using the design, final domain, and weight appropriate to each analysis; ear-specific diagnostics are in Supplement Section S8b.

Analyses incorporated NHANES survey weights, strata, and primary sampling units (PSUs). Variances were estimated by Taylor series linearization. The survey package’s adjustment was used for strata with a single PSU and for strata with only one PSU contributing observations within an analysis domain (Supplement Section S6).

In H1, stage was categorical and adjusted mean bilateral PTA4 was standardized to the weighted covariate distribution of the final H1 domain (Supplement Section S6). The three planned pairwise contrasts described above were specified before outcome analysis and formed one Holm multiplicity family (Holm 1979). They estimate adjusted between-stage PTA4 differences, not effects of individual stage-defining components.

We examined the ordering of the adjusted stage means descriptively and assessed between-stage differences using the five-stage omnibus Wald test and the three planned pairwise comparisons among Stages 2-4. The omnibus tested whether the covariate-adjusted mean bilateral PTA4 thresholds differed across the five stages; it did not test whether the means increased in order. Stage 0 contained a single participant and had no nominal stage-specific design degrees of freedom, so the omnibus was interpreted cautiously. The three planned pairwise comparisons among Stages 2-4 had much stronger survey-design support.

The glycemic staging algorithm was corrected after earlier hearing results had been reviewed. The timing and inferential implications of the glycemic staging correction are described in Supplement Section S1.

Multiplicity adjustment followed the analytic structure of each hypothesis. The nine H2 component tests formed one Holm multiplicity family. For H3, the six post-hoc Stage 2-4 interaction tests formed a separate Holm multiplicity family. For H4, the six joint tests of stage-specific coefficients formed one Holm multiplicity family, and the five within-stage block tests (two hypertension and three adiposity tests) formed another. Secondary kidney and lipid phenotype tests retained unadjusted p values only.

All hypothesis tests used design degrees of freedom (df) for the corresponding final survey domain. Reported df are denominator design df; numerator df depended on the coefficient set tested. All reported confidence intervals (CIs) were pointwise 95% intervals. Analyses used R 4.6.1 (R Core Team 2026) and survey 4.5 (Lumley 2026).

## 3 Results

### 3.1 Stage ascertainment and primary analytic sample

In the full age 70-79 domain (n = 994), the unweighted counts were 1 participant in Stage 0, 19 in Stage 1, 212 in Stage 2, 192 in Stage 3, and 279 in Stage 4; 291 (29.3%) remained unresolved. In the positive fasting-weight age domain (n = 393), 307 had resolved stage, 357 had bilateral PTA4, 390 had complete primary covariates, and 285 met all H1 requirements. These counts describe overlapping groups; they are not successive exclusions.

The H1 analytic sample included 1, 18, 103, 65, and 98 participants in Stages 0-4, respectively. Across the H1 design there were 24 strata and 49 nested PSUs, giving denominator design df = 25. The nominal design degrees of freedom within Stages 0-4 were 0, 2, 18, 12, and 20, respectively. Selected characteristics of the H1 analytic sample are shown in Table 2.

**Table 2.**
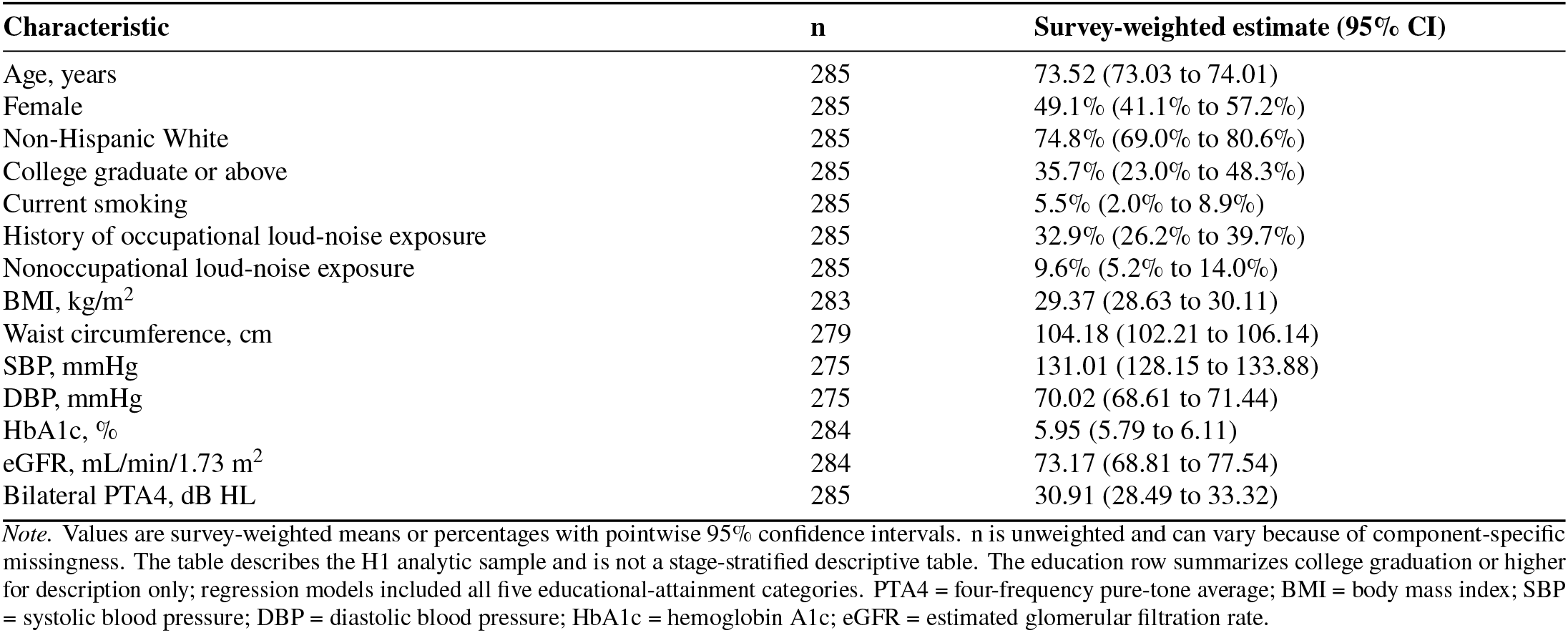
Selected characteristics of the H1 analytic sample.

| Characteristic | n | Survey-weighted estimate (95% CI) |
| --- | --- | --- |
| Age, years | 285 | 73.52 (73.03 to 74.01) |
| Female | 285 | 49.1% (41.1% to 57.2%) |
| Non-Hispanic White | 285 | 74.8% (69.0% to 80.6%) |
| College graduate or above | 285 | 35.7% (23.0% to 48.3%) |
| Current smoking | 285 | 5.5% (2.0% to 8.9%) |
| History of occupational loud-noise exposure | 285 | 32.9% (26.2% to 39.7%) |
| Nonoccupational loud-noise exposure | 285 | 9.6% (5.2% to 14.0%) |
| BMI, kg/m<sup>2</sup> | 283 | 29.37 (28.63 to 30.11) |
| Waist circumference, cm | 279 | 104.18 (102.21 to 106.14) |
| SBP, mmHg | 275 | 131.01 (128.15 to 133.88) |
| DBP, mmHg | 275 | 70.02 (68.61 to 71.44) |
| HbA1c, % | 284 | 5.95 (5.79 to 6.11) |
| eGFR, mL/min/1.73 m<sup>2</sup> | 284 | 73.17 (68.81 to 77.54) |
| Bilateral PTA4, dB HL | 285 | 30.91 (28.49 to 33.32) |

*Note.* Values are survey-weighted means or percentages with pointwise 95% confidence intervals. n is unweighted and can vary because of component-specific missingness. The table describes the H1 analytic sample and is not a stage-stratified descriptive table. The education row summarizes college graduation or higher for description only; regression models included all five educational-attainment categories. PTA4 = four-frequency pure-tone average; BMI = body mass index; SBP = systolic blood pressure; DBP = diastolic blood pressure; HbA1c = hemoglobin A1c; eGFR = estimated glomerular filtration rate.

### 3.2 Operational CKM stage and bilateral PTA4

Adjusted bilateral PTA4 means from the five-stage model were 9.49, 24.3, 31.0, 29.0, and 33.4 dB HL for Stages 0-4. The Stage 0 point estimate is retained for completeness; its model-based CI is omitted from the main display because Stage 0 consists of one participant in one PSU with nominal stage-specific design df = 0. The five-stage omnibus Wald statistic, part of the primary H1 analysis, was F(4, 25) = 21.4, p = 9.01 × 10^−8^. Adjusted stage means and the planned pairwise comparisons are summarized in Table 3 and Figure 1.

**Table 3.**
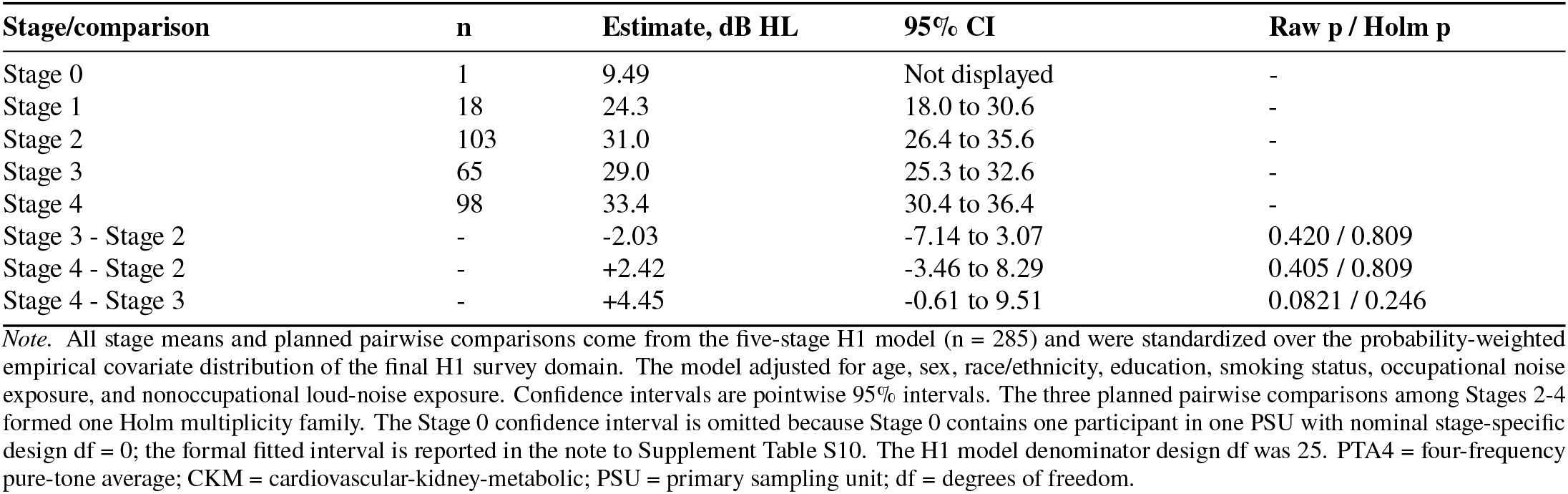
Five-stage H1 model: adjusted bilateral PTA4 and planned pairwise comparisons among Stages 2-4.

**Figure 1.**
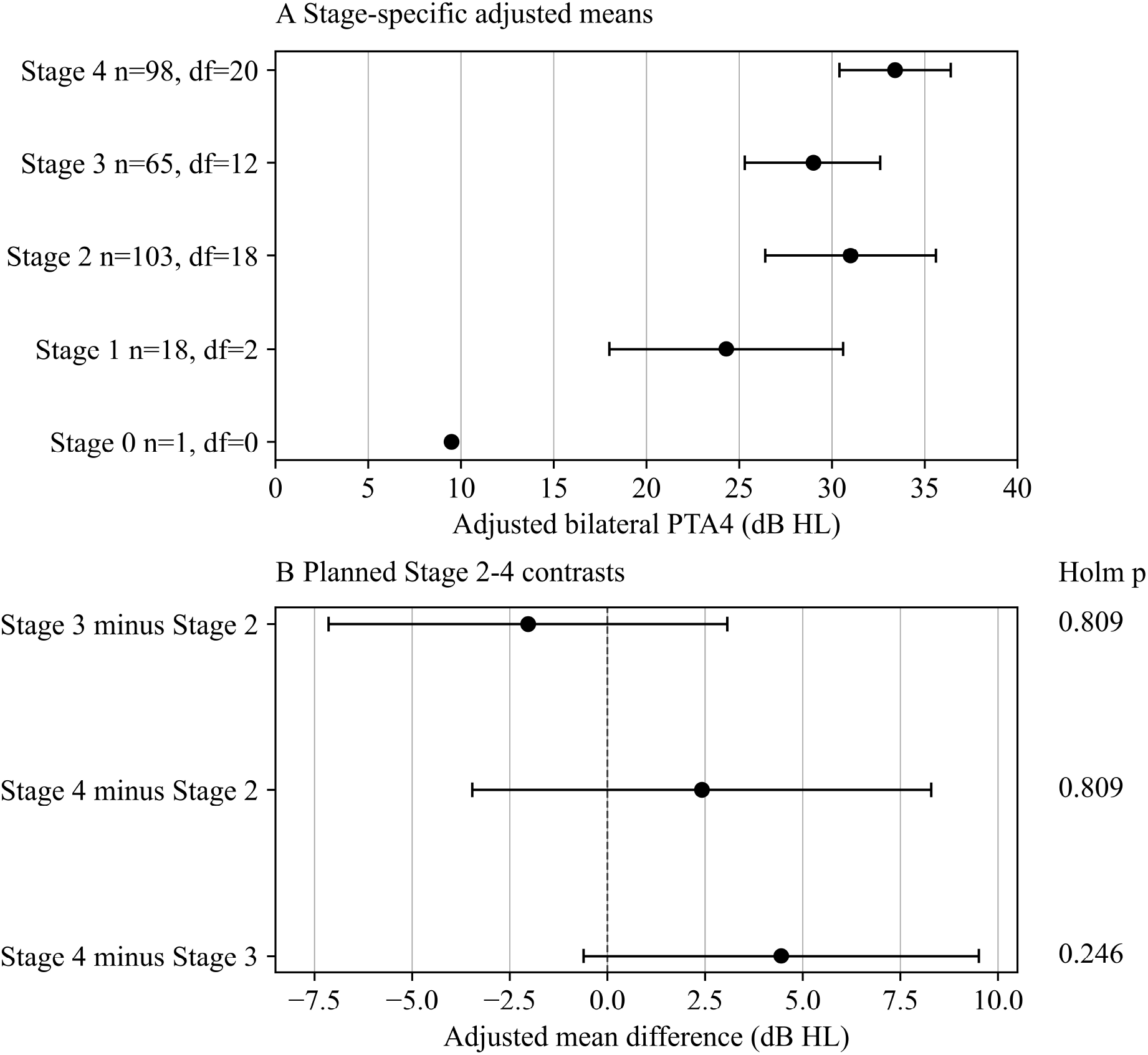
Adjusted bilateral PTA4 from the five-stage H1 model. (A) Adjusted means by operational cardiovascular-kidney-metabolic (CKM) stage, with unweighted n and nominal stage-specific design-support degrees of freedom (df). Pointwise 95% CIs are shown for Stages 1-4 and use the H1 denominator design df of 25; Stage 0 is shown without an interval because it contains one participant in one primary sampling unit, with its formal fitted interval reported in Supplement Table S10. (B) The three planned pairwise contrasts among Stages 2-4 (Stage 3 - Stage 2, Stage 4 - Stage 2, Stage 4 - Stage 3), with pointwise 95% CIs and Holm-adjusted p values. The dashed line marks zero; none met the Holm criterion. CIs are not multiplicity-adjusted. PTA4 denotes the bilateral four-frequency pure-tone average at 0.5, 1, 2, and 4 kHz.

None of the three planned pairwise comparisons among Stages 2, 3, and 4 met the Holm-adjusted significance criterion. Stage 4 minus Stage 3 was +4.45 dB HL (95% CI, − 0.61 to 9.51; raw p = 0.0821; Holm p = 0.246). A separate post-hoc Stage 2-4 refit (n = 266) gave F(2, 25) = 1.74, p = 0.197; adjusted mean bilateral PTA4 thresholds were 31.16, 28.95, and 33.31 dB HL for Stages 2, 3, and 4.

Robustness analyses gave F(3, 25) = 4.10, p = 0.0169 after collapsing Stages 0/1 and F(3, 25) = 2.77, p = 0.0626 in a separate Stage 1-4 refit (n = 284). The collapsed model retained the Stage 0 observation within a 19-person group, and the two models answer different questions; their p values do not directly isolate Stage 0 influence. Exact coefficient-block diagnostics are in Supplement Table S13.

### 3.3 Individual CKM-related measures

Associations for the nine CKM-related components are summarized in Table 4.

**Table 4.**
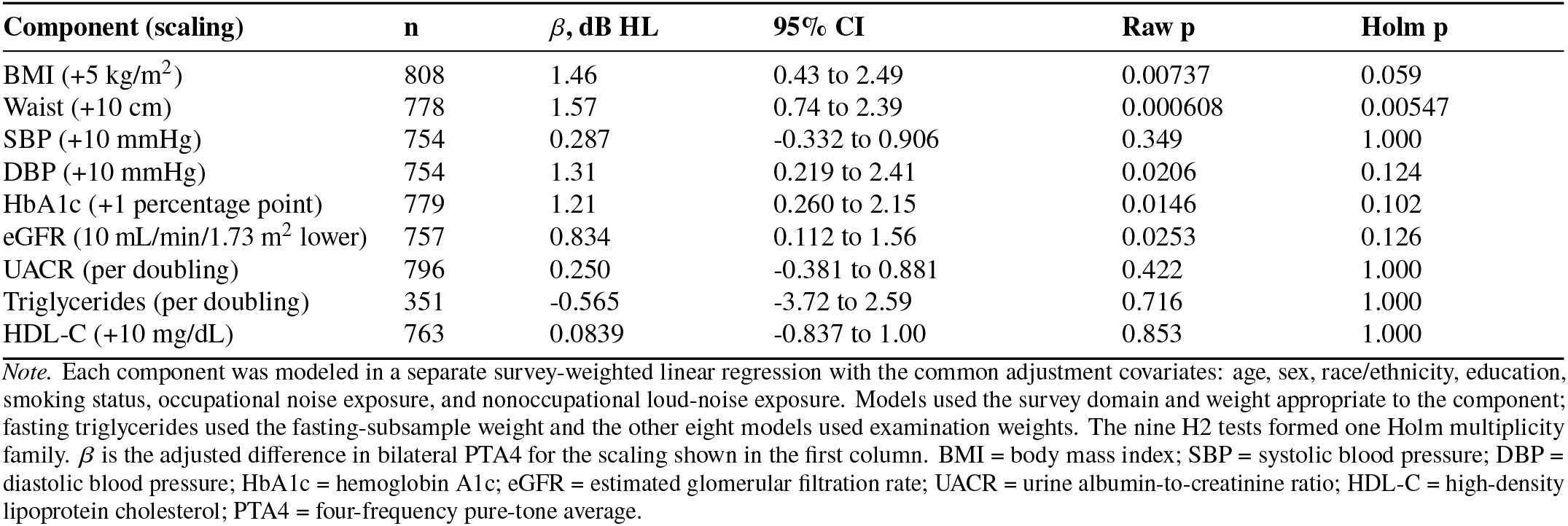
Associations of continuous CKM-related components with bilateral PTA4.

Waist circumference was the only H2 component that met the nine-test Holm criterion. In the main waist-circumference model, a 10-cm greater waist circumference corresponded to 1.57 dB HL higher bilateral PTA4 (95% CI, 0.74 to 2.39; Holm p = 0.00547; n = 778). The estimate was 1.62 dB HL per 10 cm after excluding PTA4 top-codes (95% CI, 0.88 to 2.37; p = 0.000142; n = 752). A natural-spline diagnostic did not provide clear evidence of nonlinearity, F(2, 25) = 2.23, p = 0.128.

### 3.4 Stage-by-component interactions

The full five-stage interaction analysis was structurally non-estimable because Stage 0 contained one participant. In the post-hoc Stage 2-4 restriction, none of six interaction tests met the Holm-adjusted significance criterion. The waist interaction had raw p = 0.0140 and Holm p = 0.0840; Holm p values for SBP, HbA1c, eGFR, UACR, and HDL-C were 0.276, 0.892, 1.000, 1.000, and 0.570. These results do not establish equality of slopes across stages.

### 3.5 Post-hoc within-stage analyses

Within the two exploratory H4 families, one of six joint tests of stage-specific coefficients and one of five within-stage block tests met their respective Holm criteria; both involved adiposity-related models. In the paired waist and 10-year weight-change model, the joint test of the stage-specific waist coefficients gave F(3, 25) = 22.3, Holm p = 1.83 × 10^−6^. Per 10-cm greater waist circumference, the estimated PTA4 differences were 5.09 dB HL in Stage 2 (95% CI, 3.42 to 6.77), 0.91 dB HL in Stage 3 (− 1.40 to 3.21), and 2.60 dB HL in Stage 4 (− 1.05 to 6.25). These pointwise intervals describe the stage-specific estimates and do not test differences between stages. The Stage 2 adiposity block test gave F(3, 17) = 9.76, Holm p = 0.00283; the other within-stage block tests did not meet the Holm criterion. The hypertension-profile contrasts had wide confidence intervals, leaving the size and direction of the differences uncertain. Full H4 results are reported in Supplement Tables S18-S24.

## 4 Discussion

This study did not establish operational CKM stage as a graded indicator of bilateral PTA4 in adults aged 70-79 years. The distinction between CKM stage and audiometric hearing sensitivity is central to interpreting this finding. The CKM hierarchy orders combinations of cardiovascular, kidney, and metabolic abnormalities, and people assigned to the same stage can have different clinical profiles (Ndumele et al. 2026). Its ordering therefore does not specify how hearing thresholds should vary across stages. The present findings leave that relationship unresolved. Although variation in component profiles provides a reason to examine individual measures alongside stage, this study did not determine whether such variation accounted for stage means that did not increase consistently.

The precision and stage coverage of this study constrain this interpretation. Sparse support for Stages 0 and 1 limited the assessment of hearing among participants meeting the criteria for those stages. The later-stage contrasts remained compatible with a range of hearing differences. The significant five-stage omnibus provided evidence of heterogeneity among the adjusted means, without establishing their ordering or identifying which stage differences accounted for that heterogeneity. The supplementary post-hoc Stage 2-4 refit did not provide evidence of heterogeneity among those stages. Neither that result nor the planned contrasts demonstrated equivalent thresholds, and the refit did not test directional ordering. The evidence therefore remains insufficient to characterize a graded stage-hearing relationship.

A prior longitudinal CKM study addressed the onset of perceived hearing difficulty. In Chinese adults aged ≥ 45 years, Stage 4 was associated with higher odds of incident self-reported hearing impairment than Stage 0 (Zhang et al. 2026). The present study assessed measured hearing thresholds at a single examination. These outcomes capture different aspects of hearing: perceived difficulty and audiometric sensitivity are related but are not interchangeable (de Gruy et al. 2023). The stage definitions also differed. The Chinese study defined Stage 3 by eGFR 30-59 mL/min/1.73 m^2^ without established cardiovascular disease, whereas the present study used very-high kidney risk or PREVENT 10-year total cardiovascular disease risk ≥ 20%, in the absence of observed clinical cardiovascular disease (Zhang et al. 2026). Our findings consequently do not provide a direct replication or refutation of the longitudinal association. These differences limit comparison without establishing the reason for differing results.

Evidence concerning change in cardiovascular health further distinguishes the question addressed here. Improvement in Life’s Essential 8 was associated with lower odds of hearing loss under four-frequency and high-frequency better-ear definitions, while the association did not reach statistical significance under the three-frequency definition (Yévenes-Briones et al. 2026). That exposure describes change in a cardiovascular-health score, which differs from membership in a CKM stage defined by clinical and risk criteria. The study therefore provides evidence relevant to cardiovascular health and hearing, but it does not establish that hearing thresholds increase across the CKM hierarchy. Nor does the difference in statistical significance across hearing definitions demonstrate that the corresponding associations differed from one another.

The waist association offers a more specific connection with the existing adiposity literature. Greater waist circumference was associated with higher bilateral PTA4, consistent in direction with the association reported for measured hearing sensitivity in the HUNT study (Engdahl et al. 2015). Larger waist circumference was also associated with greater risk of self-reported hearing loss in a prospective cohort of women (Curhan et al. 2013). The present study contributes an estimate for continuous bilateral PTA4 in adults aged 70-79 years, although differences in population and outcome measurement prevent treating these studies as estimates of the same relationship. Proposed metabolic and vascular pathways linking obesity with auditory dysfunction provide plausible mechanistic context, but their contribution remains uncertain and was not tested in this analysis (Terreros H., Munoz, and D’Espessailles Tapia 2025). The waist estimate was similar after excluding top-coded thresholds. Its clinical importance for an individual remains uncertain, and the association does not establish a hearing benefit from reducing waist circumference.

An association with waist circumference can coexist with uncertain stage comparisons because stage assignment and waist circumference represent different exposures. Waist circumference varies within stage groups, while stage membership reflects combinations of criteria. The analyses also used different survey domains and weights. These distinctions preclude interpreting the waist result as evidence that waist circumference is a better hearing marker than CKM stage. H2 consisted of separate component models and did not test whether waist circumference added information beyond stage or the other components. Its position as the only H2 measure meeting the nine-test Holm criterion should therefore not be used to rank the components or to explain the stage pattern.

The blood-pressure comparisons illustrate why the remaining component results require similarly specific interpretation. An association between hypertension and hearing loss, including high-frequency involvement, has been reported (Huang et al. 2025). Persistent hypertension was associated with poorer speech-in-noise performance in another study, whereas its association with audiometric thresholds was not statistically significant (Ting et al. 2022). A Finnish cohort did not detect a statistically significant association between cardiovascular disease and deterioration in better-ear hearing thresholds over 13 years (Lohi et al. 2022). Current SBP and DBP, a history of persistent hypertension, and clinical cardiovascular disease capture different aspects of cardiovascular health. Speech-in-noise performance and pure-tone thresholds also assess different hearing functions. The present estimates concern current blood pressure and bilateral PTA4; their failure to meet the Holm criterion does not settle the broader relationship between cardiovascular health and hearing.

H3 examined whether component-PTA4 associations differed across stages, whereas H4 examined associations between selected within-stage phenotypes and PTA4. H3 did not establish differences in component slopes across Stages 2-4 after Holm correction, leaving stage-related variation unresolved. Within the two H4 families, one of six parameter-level tests and one of five multidimensional stage-specific tests met their respective Holm criteria. Both involved adiposity-related models and overlapped the H2 waist analysis. These findings provide additional exploratory description of adiposity-related associations within the study but do not provide independent replication. In particular, the waist parameter-level test did not compare slopes between stages, and the Stage 2 adiposity block concerned several characteristics jointly. Neither result establishes a stronger waist association in Stage 2 or identifies a single feature responsible for the block association. Because H4 was developed after hearing-outcome review, these findings remain hypothesis-generating.

The hypertension-profile contrasts describe combinations of characteristics. Paired representative profiles differed simultaneously in hypertension duration, current SBP, medication-class composition, and current-treatment duration. The contrasts therefore cannot be attributed to a single feature, while their wide confidence intervals indicate uncertainty about the joint profile differences in PTA4. Medication-class composition remains an observed phenotype marker, not a measure of treatment intensity, hypertension severity, or resistant hypertension.

Longitudinal studies with fuller CKM ascertainment and adequate representation across stages could examine whether changes in stage and individual components accompany changes in audiometric thresholds.

### 4.1 Limitations

The cross-sectional design limits interpretation of temporal ordering and causality. CKM stage and hearing thresholds were measured at the same examination, whereas age-related hearing loss can develop over many years. Current CKM stage may therefore not represent cardiovascular, kidney, and metabolic status during the period in which hearing thresholds worsened. Adjustment for the measured covariates cannot resolve this limitation. Residual confounding also remains possible, including from differences in the intensity and duration of noise exposure.

A second limitation is that CKM stage was operationalized from ascertainable NHANES domains rather than complete clinical CKM assessment. Some domains were unavailable or incomplete, kidney risk reflected a single examination, and operational diabetes did not distinguish type 1 from type 2 diabetes. PREVENT used diagnosed-diabetes history rather than the broader laboratory-inclusive staging definition. Stage-classification uncertainty may therefore have affected estimated between-stage differences.

A third limitation concerns hearing-outcome measurement. Air-conduction audiometry cannot distinguish sensorineural, conductive, and mixed loss; bilateral PTA4 may obscure ear- or frequency-specific patterns; and code 666 was assigned 126 dB HL as an analytic no-response convention rather than representing a measured threshold.

Fourth, unresolved staging and complete-case requirements resulted in substantial analytic selection. Stage remained unresolved for 291 of 994 participants in the unweighted target-age sample. Within the positive fasting-weight domain, 285 of 393 participants met all H1 requirements for resolved stage, bilateral PTA4, and complete common covariates. Included and excluded participants differed in observed characteristics (Supplement Table S9). Among 80 participants with unresolved Stage 3 evidence in this domain, 43 lacked a required PREVENT input and 37 had complete required PREVENT inputs but at least one continuous value outside the accepted range (Supplement Section S7; Table S8). NHANES survey weights do not necessarily correct for these study-specific inclusion requirements. Selection may therefore have biased the estimated stage differences as well as limiting their generalizability, although the available data cannot determine the direction or magnitude of that bias.

Fifth, full-hierarchy inference was limited by sparse support for Stages 0 and 1: H1 included one Stage 0 and 18 Stage 1 participants. The five-stage omnibus detects any difference among means, not a progressive gradient. Its statistic changed substantially after correction of the glycemic staging algorithm (Supplement Section S8). Sparse support for Stages 0 and 1 is a separate structural limitation of the five-stage comparison. Because historical hearing-outcome results had already been viewed before correction of the glycemic staging algorithm, the corrected H1 analysis was not outcome-blinded and is not interpreted as a strictly prospective confirmatory reanalysis; the correction chronology is detailed in Supplement Section S1. The planned pairwise comparisons among Stages 2-4 remained imprecise.

Finally, the H4 analyses were developed after review of the hearing results and remain hypothesis-generating. The stage-specific multidimensional hypertension models included only 28 or 29 participants, with six design degrees of freedom and 19 or 20 model-matrix columns. The available survey-design information was limited relative to model complexity, making the resulting estimates imprecise and potentially unstable. Representative hypertension profiles differed simultaneously in hypertension duration, current SBP, medication-class composition, and current-treatment duration, so their contrasts cannot be attributed to any single feature. These H4 findings require independent evaluation rather than confirmatory interpretation.

## 5 Conclusion

Adjusted mean PTA4 did not increase with each successive CKM stage. The small numbers of participants in Stages 0 and 1 limited comparisons across the full hierarchy, and the planned Stage 2-4 comparisons remained imprecise. These data do not establish operational CKM stage as a graded indicator of PTA4 in adults aged 70-79 years. Greater waist circumference was associated with higher bilateral PTA4, although the clinical importance of the estimated difference remains uncertain.

## Data Availability

NHANES 2017-March 2020 pre-pandemic public-use data, documentation, and codebooks are freely available from the U.S. Centers for Disease Control and Prevention, National Center for Health Statistics. Study-planning materials are available on the Open Science Framework at DOI: 10.17605/OSF.IO/BCNV7. The complete analytic archive was not publicly deposited at the time this manuscript was prepared.

https://wwwn.cdc.gov/nchs/nhanes/search/datapage.aspx?Component=Demographics&Cycle=2017-2020

https://wwwn.cdc.gov/nchs/nhanes/search/datapage.aspx?Component=Examination&Cycle=2017-2020

https://wwwn.cdc.gov/nchs/nhanes/search/datapage.aspx?Component=Laboratory&Cycle=2017-2020

https://wwwn.cdc.gov/nchs/nhanes/search/datapage.aspx?Component=Questionnaire&Cycle=2017-2020

## Author contributions (CRediT)

Dinh Minh Le: Conceptualization, Methodology, Software, Validation, Formal analysis, Investigation, Data curation, Visualization, Project administration, Writing - original draft, Writing - review & editing.

Son Truc Tran: Writing - review & editing.

Van Chien Nguyen: Writing - review & editing.

## Data availability statement

NHANES 2017-March 2020 pre-pandemic public-use data and documentation are available from the National Center for Health Statistics. A study-planning record is available on the Open Science Framework (doi:10.17605/OSF.IO/BCNV7). The corresponding author can supply versioned analysis code and audit materials upon reasonable request. The complete reproducibility archive had no permanent public DOI at manuscript preparation.

## Ethical approval and informed consent

NHANES protocols were reviewed and approved by the National Center for Health Statistics Ethics Review Board. NHANES 2017-2018 was covered by Protocol #2018-01, with continuation of Protocol #2011-17 through October 26, 2017; NHANES 2019-2020 continued Protocol #2018-01. NHANES obtained informed consent for the original interview and examination data collection. No new consent was obtained for this deidentified secondary analysis.

## Use of generative artificial intelligence

OpenAI ChatGPT (GPT-5.6 Sol) and Anthropic Claude (Opus 5.0) assisted only with language refinement, structural editing, and source and citation checking during later manuscript revision. The authors developed the scientific question, study design, and initial manuscript draft, prepared the data, and performed the statistical analyses, including developing the R code used to produce the reported results, without generative-AI assistance. The authors reviewed both tools’ applicable terms of use and confirmed that their use was consistent with those terms and suitable for publication. They verified all AI-assisted revisions for accuracy and originality, checked the references, and take full responsibility for the submitted work.

