## Supplementary Material for "Audiometric hearing thresholds and operational cardiovascular-kidney-metabolic staging in U.S. adults aged 70-79 years: a cross-sectional NHANES analysis"

#### S1. Analysis chronology and inferential status

The original H1-H3 analytic questions and the three planned H1 Stage 2-4 contrasts preceded outcome analysis. Historical hearing-outcome results had already been viewed when an internal audit identified that design-valid fasting plasma glucose had not been used as an independent Stage 1 prediabetes or Stage 2 diabetes criterion in the operational implementation of the 2026 CKM guideline (Ndumele et al. 2026). The glycemic staging algorithm was corrected and H1-H3 were rerun. Because historical hearing-outcome results had already been viewed before correction of the glycemic staging algorithm, the corrected H1 analysis was not outcome-blinded and is not interpreted as a strictly prospective confirmatory reanalysis. The final Stage 2-4 H3 interaction refits were conducted post-hoc after the original five-stage H3 models proved structurally non-estimable. H4 was developed later, after outcome results were available, and is explicitly post-hoc. The corrected-stage H4 rerun reused the previously defined phenotype variables, covariates, survey design, feasibility rules, model forms, profile registry, and multiplicity families.

**Table S1. Analysis chronology and inferential status**

| Phase | Outcome information available? | Role in final report |
|---|---|---|
| Study planning | No final hearing results used | Defined H1-H3, bilateral PTA4, covariate domains, and survey-regression framework. |
| Historical H1-H3 analysis | Yes, after models were run | Revealed initial hearing results. |
| Glycemic staging correction | Historical hearing results already known | Corrected exposure implementation; H1-H3 rerun. |
| H4 development | Hearing results already known | Post-hoc exploratory within-stage phenotype program. |
| Corrected-stage H4 rerun | H4 results already known | Correctness/provenance check; no new phenotype definition, contrast, or multiplicity family. |

#### S2. Operational CKM stage hierarchy and tri-state logic

Final CKM stage was assigned as Stage 4 > Stage 3 > Stage 2 > Stage 1 > Stage 0, consistent with the CKM staging framework and its subsequent multisociety guideline (Ndumele et al. 2026). Three-state logic preserved positive, negative, and unresolved evidence, so unresolved evidence for a higher stage could prevent assignment to a lower stage. Stage 0 was not a residual category: it required resolved absence of Stages 1-4 together with explicit normal Stage 0 evidence, including no excess adiposity, normal glycemia, normal blood-pressure/treatment evidence, normal lipid evidence including no low HDL-C, no moderate or higher kidney risk, no Stage 3 evidence, and no observed clinical cardiovascular disease under the operational rules. Guideline domains not comprehensively ascertained by the sources used in this study were excluded from the operational construct. Within the implemented domains, missing required inputs or values outside the accepted calculator ranges were retained as unresolved and could prevent assignment to a lower stage. Stage-defining variables were not multiply imputed.

**Table S2. Operational CKM stage hierarchy**

| Stage | Operational rule in this study |
|---|---|
| 0 | Resolved absence of Stages 1-4 plus explicit normal Stage 0 evidence; not a residual category. |
| 1 | Excess adiposity or prediabetes, with higher-stage criteria resolved as absent. |
| 2 | Hypertension, hypertriglyceridemia, metabolic syndrome, operational diabetes, or moderate or higher kidney risk. |
| 3 | Very-high kidney risk or PREVENT 10-year total CVD risk ≥20%, without observed clinical CVD. |
| 4 | Observed clinical CVD in an operational CKM context. |

In the full age 70-79 domain (n = 994), the corrected algorithm classified 1/19/212/192/279 participants as Stages 0/1/2/3/4, respectively; 291 remained indeterminate. Operational CKM context for Stage 4 was satisfied by definite evidence of any of the following: excess adiposity; dysglycemia from diabetes history, HbA1c in the prediabetes/diabetes range, or design-valid fasting plasma glucose in the prediabetes/diabetes range; hypertension; hypertriglyceridemia; metabolic syndrome; or moderate or higher kidney risk. Stage 4 then required observed clinical cardiovascular disease together with definite CKM context. The 291 indeterminate classifications are an unweighted descriptive count in the full age domain and are part of the three-state exposure definition; they are not a population prevalence estimate of staging failure. Unresolved higher-stage evidence was not recoded as absence. The 2026 guideline further subdivides clinical Stage 4 according to kidney-failure status (Stages 4a and 4b) (Ndumele et al. 2026). This study did not retain that subdivision and analyzed the operational Stage 4 group as a single category.

#### S3. Source-to-operationalization map

Table 4 of the 2026 CKM guideline specifies the Stage 1 BMI and waist-circumference thresholds used here, including BMI ≥23 kg/m$^2$ and waist circumference ≥80 cm in women or ≥90 cm in men for Asian ancestry (Ndumele et al. 2026). The operational Stage 1 adiposity rule uses the BMI criterion OR the waist criterion. The operational diabetes definition combined clinician-diagnosed diabetes history with HbA1c and design-valid fasting plasma glucose criteria and did not distinguish type 1 from type 2 diabetes.

**Table S3. Source-to-operationalization map**

| Construct | NHANES source / input | Operational definition | Missing-data handling / limitation |
|---|---|---|---|
| BMI | BMXBMI | Standard ≥25 kg/m$^2$; Asian ≥23 kg/m$^2$; BMI OR waist criterion. | Missing BMI remained unknown. |
| Waist circumference | BMXWAIST | Standard ≥102 cm men / ≥88 cm women; Asian ≥90 / ≥80 cm; waist OR BMI criterion. | Missing waist remained unknown. |
| Measured BP | BPXOSY1-3; BPXODI1-3 | Mean SBP/DBP retained with ≥2 valid readings. | <2 valid readings left measured-pressure evidence unresolved. |

| Construct | NHANES source / input | Operational definition | Missing-data handling / limitation |
|---|---|---|---|
| Current antihypertensive treatment | BPQ050A + history | Positive with current prescribed treatment. | Unresolved questionnaire combinations remained unknown. |
| HbA1c | LBXGH | Diabetes ≥6.5%; prediabetes 5.7%-6.4% after diabetes exclusion. | Observed valid laboratory value required. |
| Fasting plasma glucose | LBXGLU + WTSAFPRP>0 | Diabetes ≥126 mg/dL; prediabetes 100 to <126 mg/dL after diabetes exclusion. | Without positive fasting weight, FPG evidence remained unknown. |
| Fasting triglycerides | LBXTR + WTSAFPRP>0 | Hypertriglyceridemia ≥150 mg/dL. | Without design-valid fasting data, TG evidence remained unknown. |
| HDL-C | LBDHDD | Low HDL-C <40 mg/dL men or <50 mg/dL women for metabolic-syndrome scoring. | Missing HDL-C remained unknown. |
| Metabolic syndrome | Waist, HDL-C, TG, BP/treatment, FPG | True when ≥3 of 5 criteria definitely positive. | False only when positive + unknown could not reach 3; otherwise unknown. |
| Kidney function | Creatinine eGFR + UACR | 2021 CKD-EPI creatinine eGFR without race; KDIGO risk regions. | Single visit does not establish chronicity. |
| PREVENT Stage 3 risk | AHAprevent 1.0.0; base total-CVD | 10-y total CVD risk ≥20%; dm=clinician diabetes history; current BP treatment; statin use. | Missing/out-of-range required input or observed CVD prevented scoring. |
| Observed clinical CVD | MCQ160b/c/d/e/f | Heart failure, CHD, angina, MI, or stroke; positive if any was positive. | PAD and AF were not comprehensively ascertained by the sources used in staging and were excluded; direct subclinical CVD measures were also excluded. Exclusion was not coded as absence. |
| Audiometry top-code | P_AUX code 666 | Assigned 126 dB HL for quantitative analysis. | Analytic top-code for no-response threshold. |
| Bilateral PTA4 | P_AUX 0.5,1,2,4 kHz | Mean of valid right- and left-ear PTA4; both ears required. | 1-kHz reliability and retest rules applied before PTA4. |

| Construct | NHANES source / input | Operational definition | Missing-data handling / limitation |
|---|---|---|---|
| Clinician-diagnosed diabetes history (PREVENT input) | P_DIQ: DIQ010 | Yes (1) = positive history; No (2) or Borderline (3) = negative history. | Refused (7), Don't know (9), or missing = unresolved. This variable supplied the PREVENT diabetes input and was distinct from the laboratory-inclusive operational diabetes definition used for CKM staging. |

### S4. Additional implementation details for CKM components

#### 0.1 S4.1 Blood pressure and treatment

Oscillometric SBP and DBP used BPXOSY1-3 and BPXODI1-3. The arithmetic mean was calculated only with at least two valid measurements. Hypertension history used BPQ020, age at first diagnosis used BPD035, and current prescribed treatment used BPQ050A. Prescription-medication files were used for H4 medication-class composition variables, not to redefine the H1-H3 hypertension criterion.

#### 0.2 S4.2 Fasting lipids and metabolic syndrome

Design-valid fasting triglycerides required an observed LBXTR and positive fasting-subsample weight. Metabolic syndrome used abdominal obesity, low HDL-C, fasting triglycerides ≥150 mg/dL, SBP ≥130 mmHg or DBP ≥80 mmHg or current antihypertensive treatment, and design-valid fasting glucose ≥100 mg/dL (Ndumele et al. 2026). Three-state logic preserved uncertainty when the five-criterion rule could not be resolved.

#### 0.3 S4.3 Kidney risk

Creatinine-based eGFR used the 2021 CKD-EPI creatinine equation without race (Inker et al. 2021). eGFR and UACR categories and the operational risk mapping followed KDIGO categories (Kidney Disease: Improving Global Outcomes (KDIGO) CKD Work Group 2024): eGFR categories were G1 ≥90, G2 60 to <90, G3a 45 to <60, G3b 30 to <45, G4 15 to <30, and G5 <15 mL/min/1.73 $m^2$; UACR categories were A1 <30, A2 30 to <300, and A3 ≥300 mg/g. The operational mapping treated G1/G2-A1 as low risk; G1/G2-A2/A3, G3a-A1/A2, and G3b-A1 as moderate-to-high risk; and G3a-A3, G3b-A2/A3, and all G4/G5 states as very high risk.

#### 0.4 S4.4 PREVENT

PREVENT risk was calculated with AHAprevent version 1.0.0 using the base total-CVD implementation (Khan et al. 2024; American Heart Association 2026). The required inputs were sex, age, total cholesterol, HDL-C, SBP, BMI, eGFR, diabetes, smoking, blood-pressure treatment, and statin use (Khan et al. 2024). For PREVENT, the required binary diabetes input was operationalized using clinician-diagnosed diabetes history. The broader laboratory-inclusive diabetes definition was used elsewhere in CKM staging. The use of diabetes history was an implementation choice and could affect Stage 3 ascertainment relative to substituting the broader operational diabetes definition. Blood-pressure treatment represented current prescribed antihypertensive treatment, and the statin input represented statin use rather than all lipid-lowering therapy. For this implementation, accepted continuous input ranges were age 30-79 years, SBP 90-200 mmHg, total cholesterol 130-320 mg/dL, HDL-C 20-100 mg/dL, eGFR 15-140 mL/min/1.73 $m^2$, and BMI 18.5-39.9 kg/$m^2$ (American Heart Association 2026). These were implementation validity limits rather than biological risk cut points. A required missing or out-of-range input, or observed established cardiovascular disease, prevented primary-prevention risk calculation. Age, sex, and smoking are PREVENT inputs and were also common H1 adjustment covariates; adjusted stage comparisons therefore condition on some information used in staging.

##### 0.5 S4.5 Diagnosis-age handling for H4 disease-duration variables

Hypertension duration used BPD035, introduced in S4.1, and diabetes duration used DID040. BPD035 = 12 denotes age 12 years or younger, and BPD035 = 80 denotes age 80 years or older. DID040 = 666 denotes age younger than 1 year, and DID040 = 80 denotes age 80 years or older. Codes 777 and 999 denote refused and don't know, respectively, in both variables.

Exact diagnosis ages were retained within 13-79 years for BPD035 and 1-79 years for DID040. The non-exact codes described above and missing responses yielded missing exact diagnosis ages. The source-validation code stopped on unexpected raw codes or values.

Hypertension duration was calculated for participants aged 70-79 years as current age minus exact diagnosis age when hypertension history was positive and exact diagnosis age did not exceed current age. Diabetes duration was calculated as current age minus exact diagnosis age when diabetes history was positive and exact diagnosis age did not exceed current age, without an age-domain restriction at the derivation step. H4 analyses remained restricted to adults aged 70-79 years. Execution stopped if an exact diagnosis age exceeded current age. No duration was imputed.

A missing duration made a participant ineligible for the corresponding H4 phenotype core. The hypertension core required diagnosed hypertension history and nonmissing hypertension duration, current SBP, and current antihypertensive-treatment status. The diabetes core required clinician-diagnosed diabetes and nonmissing diabetes duration, HbA1c, and diabetes-treatment context. Participants with non-exact diagnosis-age responses were therefore ineligible for the corresponding parameter-level duration model.

#### S5. Audiometry rules and targeted sensitivity analyses

Air-conduction thresholds came from P_AUX (National Center for Health Statistics 2022). At routine 1 kHz, the first and second readings were treated as a reliability pair: if both were numeric and differed by ≤10 dB, the first reading supplied the standard threshold; if both were 666, 666 was retained; otherwise routine 1 kHz was unresolved. An insert/crossover retest replaced the corresponding standard threshold. Retest code 888 remained missing. Final code 666 was assigned 126 dB HL; code 888 and unresolved values remained missing. These rules were applied in the context of NHANES audiometric threshold coding and prior NHANES threshold analyses (Humes 2023).

Among 821 target-age participants with valid bilateral PTA4, 26 had at least one top-coded PTA4 component and 44 had at least one insert/crossover retest used in the final threshold set. In H1, 10 participants had a top-coded component; nine were in Stages 2-4. A tympanometry-restricted sensitivity was considered. The available archive did not allow verification that the estimate came from the corrected analysis, so no numerical tympanometry sensitivity estimate is reported.

**Table S4. Targeted audiometry sensitivities with verified corrected provenance**

| Sensitivity analysis | n | Result |
|---|---|---|
| Five-stage H1 after excluding PTA4 top-codes | 275 | $F(4, 25) = 25.443$; $p = 1.70 \times 10^{-8}$ |
| Stage 2-4 H1 after excluding PTA4 top-codes | 257 | $F(2, 25) = 2.723$; $p = 0.0851$ |

| Sensitivity analysis | n | Result |
|---|---|---|
| H2 waist after excluding PTA4 top-codes | 752 | +1.62 dB HL per 10 cm; 95% CI, 0.88 to 2.37; p = 0.000142 |

*The five-stage no-top-code result retains the singleton Stage 0 with nominal stage-specific design df = 0 and is not interpreted as stable five-stage evidence.*

#### S6. Survey design, weights, degrees of freedom, and multiplicity

Survey objects were constructed before age-domain and model-specific restrictions, consistent with the combined pre-pandemic NHANES analytic structure (Stierman et al. 2021). H1 and H3 used the fasting-subsample weight WTSAFPRP; H2 used the MEC examination weight WTMECPRP except for triglycerides, which used WTSAFPRP; H4 used WTSAFPRP. PSUs were nested within strata in the survey design (nest = TRUE). Variance estimation used Taylor linearization; survey.lonely.psu = "adjust" and survey.adjust.domain.lonely = TRUE. Under the "adjust" option, a single-PSU stratum is centered at the overall sample mean rather than its own stratum mean; the domain option applies the adjustment when a stratum has only one contributing PSU within an analysis domain. Denominator degrees of freedom followed the model-domain survey design (nested PSUs minus represented strata; survey::degf). Age entered linearly in years. Categorical covariates were sex (Female/Male), race/ethnicity (Mexican American, Other Hispanic, Non-Hispanic White, Non-Hispanic Black, Non-Hispanic Asian, Other/multiracial), education (five DMDEDUC2 categories), smoking (never/former/current), occupational noise (no loud-noise job/ever loud-noise job/never worked), and nonoccupational loud-noise exposure (no/yes). Refused, don't-know, unexpected, skipped, and genuinely missing values remained missing rather than being assigned to a reference category. Coefficient reference levels were inherited from the factor-level ordering stored in the retained analysis objects; the exact level ordering/model-matrix coding is retained in the versioned code. H1 adjusted means used survey::svypredmeans, averaging predictions over the probability-weighted empirical covariate distribution of the analytic survey domain; pointwise 95% CIs used the domain design df and the survey-model covariance matrix. No multiple imputation was used for stage-defining variables or common covariates. Common-covariate missingness was handled by model-specific complete-case restriction; unresolved stage evidence remained unresolved rather than being imputed into a stage.

**Table S5. Stage-specific design support in the corrected H1 analytic domain**

| Stage | n | Strata | Nested PSUs | Nominal stage df |
|---|---|---|---|---|
| Stage 0 | 1 | 1 | 1 | 0 |
| Stage 1 | 18 | 12 | 14 | 2 |
| Stage 2 | 103 | 24 | 42 | 18 |
| Stage 3 | 65 | 23 | 35 | 12 |
| Stage 4 | 98 | 24 | 44 | 20 |

*The full H1 design retained 24 strata, 49 nested PSUs, and denominator design df = 25.*

Multiplicity adjustment within the predefined test families used Holm's sequentially rejective procedure (Holm 1979).

**Table S6. Multiplicity families**

| Family | Tests | Multiplicity treatment |
|---|---|---|
| H1 planned contrasts | Stage 3−2, Stage 4−2, Stage 4−3 | Holm, family size 3 |
| H2 components | 9 separate continuous component models | Holm, family size 9 |

| Family | Tests | Multiplicity treatment |
|---|---|---|
| H3 Stage 2-4 interactions | Waist, SBP, HbA1c, eGFR, UACR, HDL-C | Holm, family size 6 |
| H4 parameter-level exploratory | 6 joint parameter tests | Holm, family size 6 |
| H4 multidimensional exploratory | HTN S3/S4; waist/weight-history S2/S3/S4 | Holm, family size 5 |
| H4 kidney/lipid secondary | 4 phenotype-block tests | Raw p values only |

#### S7. Participant flow and fasting-domain selection audit

The full age-domain n was 994; 917 had audiometry records and 821 had valid bilateral PTA4. The positive fasting-weight age domain contained 393 participants, of whom 307 had resolved stage and 86 had indeterminate stage. Availability counts below overlap and do not represent a sequential attrition chain.

**Table S7. Availability within the fasting age domain**

| Availability criterion | n |
|---|---|
| Positive fasting weight, age 70-79 | 393 |
| Resolved operational CKM stage | 307 |
| Observed bilateral PTA4 | 357 |
| Complete primary covariates | 390 |
| H1 intersection | 285 |

Within the fasting-weight-eligible domain (n = 393), 86 participants had indeterminate stage; 80 had unresolved Stage 3 evidence and six were unresolved for Stage 4. All 80 Stage-3-unresolved records had unresolved PREVENT Stage-3 evidence. Among those 80, 43 had at least one missing required PREVENT input and 37 had all required inputs present but at least one continuous input outside the accepted range; these two high-level categories were mutually exclusive. Nine also had unresolved very-high kidney-risk evidence.

**Table S8. Overlapping reasons for Stage-3 unresolved status**

| Audit flag | n | Counting rule |
|---|---|---|
| PREVENT Stage-3 evidence unknown | 80 | All Stage-3-unresolved records |
| ≥1 required PREVENT input missing | 43 | High-level category |
| Required inputs complete, but ≥1 continuous input out of range | 37 | High-level category |
| Very-high kidney-risk evidence also unknown | 9 | May overlap with PREVENT reasons |
| Missing SBP | 28 | Input-specific; may overlap |
| Total cholesterol out of range | 22 | Input-specific; may overlap |
| BMI out of range | 12 | Input-specific; may overlap |
| HDL-C out of range | 8 | Input-specific; may overlap |
| Raw kidney eGFR missing | 8 | Input-specific kidney flag |
| PREVENT eGFR missing | 8 | Input-specific PREVENT flag |

*The aggregate audit did not collapse out-of-range values into presumed low-versus-high directions.*

**Table S9. Descriptive comparison of participants included versus not included in H1 within the positive fasting-weight age domain**

| Characteristic | Included in H1 (n = 285) | Not included in H1 (n = 108) |
|---|---|---|
| Age, years | 73.5 (73.0 to 74.0) | 74.2 (73.4 to 75.0) |
| Female sex | 49.1% (41.1 to 57.2) | 73.0% (63.2 to 82.7) |
| Non-Hispanic White | 74.8% (69.0 to 80.6) | 63.4% (46.5 to 80.2) |
| College graduate or above | 35.7% (23.0 to 48.3) | 20.6% (9.5 to 31.6) |
| Current smoking | 5.5% (2.0 to 8.9) | 8.1% (1.0 to 15.1) |
| Ever loud-noise job | 32.9% (26.2 to 39.7) | 25.4% (15.0 to 35.7) |
| Nonoccupational loud-noise exposure | 9.6% (5.2 to 14.0) | 4.3% (0 to 8.9) |
| BMI, $kg/m^2$ | 29.4 (28.6 to 30.1) | 29.3 (27.5 to 31.1) |
| Waist circumference, cm | 104.2 (102.2 to 106.1) | 101.2 (97.7 to 104.7) |
| SBP, mmHg | 131.0 (128.1 to 133.9) | 134.3 (128.6 to 139.9) |
| HbA1c, % | 5.95 (5.79 to 6.11) | 6.02 (5.79 to 6.26) |
| eGFR, mL/min/1.73 $m^2$ | 73.2 (68.8 to 77.5) | 78.0 (74.2 to 81.8) |
| Bilateral PTA4, dB HL | 30.9 (28.5 to 33.3) | 28.2 (24.2 to 32.2)* |
| Resolved CKM stage available | 100% | 12.0% (4.8 to 19.2) |
| Bilateral PTA4 available | 100% | 76.8% (67.0 to 86.7) |
| Primary covariates complete | 100% | 98.6% (96.9 to 100) |

Values are survey-weighted descriptive estimates (95% CIs); no between-group hypothesis tests were performed. *Bilateral PTA4 among participants not included in H1 is an available-case estimate (unweighted n = 72). Because hearing data were not observed for all excluded participants, this comparison does not determine the direction or magnitude of selection bias.
Provenance: Retained H1 selection-audit output for the positive fasting-weight age domain; these descriptive estimates were not used to select or modify the H1 model.

#### S8. H1 detailed results and robustness analyses

**Table S10. Five-stage H1 model: adjusted means and planned Stage 2-4 contrasts**

| Stage / comparison | n | Estimate, dB HL | 95% CI | Raw p / Holm p |
|---|---|---|---|---|
| Stage 0 | 1 | 9.49 | Not displayed | - |
| Stage 1 | 18 | 24.3 | 18.0 to 30.6 | - |
| Stage 2 | 103 | 31.0 | 26.4 to 35.6 | - |
| Stage 3 | 65 | 29.0 | 25.3 to 32.6 | - |
| Stage 4 | 98 | 33.4 | 30.4 to 36.4 | - |
| Stage 3 − Stage 2 | - | −2.03 | −7.14 to 3.07 | 0.420 / 0.809 |
| Stage 4 − Stage 2 | - | +2.42 | −3.46 to 8.29 | 0.405 / 0.809 |
| Stage 4 − Stage 3 | - | +4.45 | −0.61 to 9.51 | 0.0821 / 0.246 |

*Corrected global Wald:* $F(4, 25) = 21.405$, $p = 9.01 \times 10^{-8}$. *Stage 0 formal fitted CI = 4.48 to 14.50 dB HL, suppressed from the main display.*

The corrected five-stage fitted means were 9.49, 24.3, 31.0, 29.0, and 33.4 dB HL for Stages 0-4. Descriptively, the fitted point estimates increased from Stage 0 through Stage 2, decreased at Stage 3, and reached their highest value at Stage 4. This sample pattern does not establish a non-monotonic population relation, and the design-support limitations of the five-stage model remain unchanged.

Historical-to-corrected comparison. Before the fasting-glucose staging correction, Stage 0 contained two participants, with an adjusted mean of 16.36 dB HL and standard error 7.47 dB. After correction, Stage 0 contained one participant, with an adjusted mean of 9.49 dB HL and standard error 2.43 dB; standard errors for Stages 1-4 were nearly unchanged. Across the same correction, the global five-stage statistic changed from F(4, 25) = 3.257, p = 0.0279, to F(4, 25) = 21.405, p = $9.01 \times 10^{-8}$. This comparison documents sensitivity of the global Wald statistic to correction of the glycemic staging algorithm. Sparse support for Stages 0 and 1, including singleton Stage 0, is a separate structural limitation of the five-stage omnibus.

**Table S11. Post-hoc Stage 2-4-only refit**

| Quantity | Estimate, dB HL | 95% CI | Holm p |
|---|---|---|---|
| Stage 2 mean | 31.16 | 26.46 to 35.86 | - |
| Stage 3 mean | 28.95 | 25.37 to 32.52 | - |
| Stage 4 mean | 33.31 | 30.31 to 36.30 | - |
| Stage 3 − Stage 2 | −2.21 | −7.25 to 2.82 | 0.747 |
| Stage 4 − Stage 2 | +2.15 | −3.76 to 8.05 | 0.747 |
| Stage 4 − Stage 3 | +4.36 | −0.56 to 9.28 | 0.241 |

*n = 266; omnibus F(2, 25) = 1.735, p = 0.197. This model was standardized over its own analytic domain; its contrasts are not the planned contrasts from the n = 285 five-stage model.*

The Stage 0/1 collapsed model gave F(3, 25) = 4.10, p = 0.0169. This model retained the Stage 0 observation within a 19-person early-stage group. A separate Stage 1-4 refit excluded the Stage 0 observation and gave n = 284, F(3, 25) = 2.77, p = 0.0626. The separate refit and within-model coefficient-block deletion can yield the same rounded F value despite being different computations.

#### 0.6 S8b. Ear-specific outcomes and exact coefficient-block diagnostics

**Table S12. Corrected ear-specific secondary H1 results**

| Outcome | n | Full five-stage F equivalent | p |
|---|---|---|---|
| Right-ear PTA4 | 287 | 14.3 | $3.27 \times 10^{-6}$ |
| Left-ear PTA4 | 285 | 31.9 | $1.75 \times 10^{-9}$ |

These are secondary diagnostics of corrected Wald behavior, not evidence of stable five-stage separation.

**Table S13. Exact coefficient-block diagnostics for corrected H1 outcomes**

| Outcome | Full Wald | Full F | Omit Stage 0 coefficient Wald | Omit Stage 0 coefficient F | Omit Stage 0 and 1 coefficients Wald | Omit Stage 0 and 1 coefficients F / p |
|---|---|---|---|---|---|---|
| Bilateral PTA4 | 85.6 | 21.4 | 8.31 | 2.77 | 3.34 | 1.67 / 0.209 |
| Right-ear PTA4 | 57.4 | 14.3 | 8.65 | 2.88 | 4.89 | 2.45 / 0.107 |
| Left-ear PTA4 | 127.6 | 31.9 | 8.01 | 2.67 | 0.979 | 0.489 / 0.619 |

*These are within-model Wald quadratic-form diagnostics from the same corrected five-stage fit. Stage 2 was the computational reference in the diagnostic model, so the tested stage coefficient block represented Stages 0, 1, 3, and 4. Removing Stage 0 from that tested block reduced the Wald quadratic form by 90.3%, 84.9%, and 93.7% for bilateral, right-ear, and left-ear PTA4; removing Stage 0 and Stage 1 reduced it by 96.1%, 91.5%, and 99.2%. These percentages describe changes in the tested Wald quadratic form, not an additive variance decomposition or causal contribution. The separate Stage 1-4 analysis reported above was a distinct refit rather than a coefficient-block deletion.*

#### S9. H2 component models and waist diagnostics

**Table S14. H2 component models**

| Component (scaling) | n | Estimate, dB HL | 95% CI | Raw p | Holm p |
|---|---|---|---|---|---|
| BMI (+5 kg/m$^2$) | 808 | 1.46 | 0.43 to 2.49 | 0.00737 | 0.059 |
| Waist (+10 cm) | 778 | 1.57 | 0.74 to 2.39 | 0.000608 | 0.00547 |
| SBP (+10 mmHg) | 754 | 0.287 | −0.332 to 0.906 | 0.349 | 1.000 |
| DBP (+10 mmHg) | 754 | 1.31 | 0.219 to 2.41 | 0.0206 | 0.124 |
| HbA1c (+1 percentage point) | 779 | 1.21 | 0.260 to 2.15 | 0.0146 | 0.102 |
| eGFR (10 mL/min/1.73 m$^2$ lower) | 757 | 0.834 | 0.112 to 1.56 | 0.0253 | 0.126 |
| UACR (per doubling) | 796 | 0.250 | −0.381 to 0.881 | 0.422 | 1.000 |
| Triglycerides (per doubling) | 351 | −0.565 | −3.72 to 2.59 | 0.716 | 1.000 |
| HDL-C (+10 mg/dL) | 763 | 0.0839 | −0.837 to 1.00 | 0.853 | 1.000 |

Waist circumference was the only H2 component meeting the nine-test Holm criterion. The no-top-code waist estimate was 1.62 dB HL per 10 cm (95% CI, 0.88 to 2.37; p = 0.000142; n = 752). A three-degree-of-freedom natural-spline model passed numerical checks; the two-degree-of-freedom nonlinearity test was F(2, 25) = 2.23, p = 0.128. Across 49 leave-one-PSU waist models, coefficients ranged from 1.34 to 1.73 dB HL per 10 cm and remained positive.

#### S10. H3 interaction estimability and Stage 2-4 results

The five-stage H3 interaction models were structurally non-estimable because Stage 0 contained one participant and could not provide a Stage-0-specific continuous slope. No five-stage stage-specific slopes were generated. The Stage 2-4 interaction refits reported below were therefore conducted post-hoc and are exploratory.

**Table S15. Stage 2-4 H3 interaction tests**

| Component | n | F(2, 25) | Raw p | Holm p |
|---|---|---|---|---|
| Waist circumference | 260 | 5.09 | 0.0140 | 0.0840 |
| SBP | 256 | 3.26 | 0.0552 | 0.276 |

| Component | n | F(2, 25) | Raw p | Holm p |
|---|---|---|---|---|
| HbA1c | 265 | 1.27 | 0.297 | 0.892 |
| eGFR | 265 | 0.340 | 0.715 | 1.000 |
| UACR | 261 | 0.230 | 0.796 | 1.000 |
| HDL-C | 265 | 2.11 | 0.142 | 0.570 |

*No interaction met the Holm-adjusted significance criterion. A significant slope within one stage and a nonsignificant slope in another is not evidence that slopes differ.*

**Table S16. Stage-specific slopes from the post-hoc Stage 2-4 H3 interaction models**

| Component | Stage 2 slope (95% CI) | Stage 3 slope (95% CI) | Stage 4 slope (95% CI) |
|---|---|---|---|
| Waist +10 cm | 5.03 (3.31 to 6.75) | 0.905 (−1.42 to 3.23) | 1.83 (−0.85 to 4.52) |
| SBP +10 mmHg | −2.03 (−3.94 to −0.12) | 0.208 (−2.05 to 2.46) | 1.09 (−1.13 to 3.32) |
| HbA1c +1 point | 2.26 (−0.42 to 4.94) | 1.78 (−1.30 to 4.85) | −1.06 (−4.41 to 2.29) |
| eGFR (10 mL/min/1.73 $m^2$ lower) | 0.109 (−2.69 to 2.91) | 0.668 (−1.38 to 2.71) | 1.32 (−0.014 to 2.65) |
| UACR doubling | −0.090 (−1.29 to 1.11) | 0.436 (−0.63 to 1.50) | 0.018 (−2.01 to 2.05) |
| HDL-C +10 mg/dL | −1.36 (−4.08 to 1.35) | 1.52 (−0.83 to 3.88) | 1.93 (0.37 to 3.49) |

Stage-specific slopes are descriptive outputs from the interaction models and were not tested as a separate multiplicity family. A slope whose individual confidence interval excludes zero in one stage and not another does not establish that the slopes differ; the interaction tests in Table S15 are the relevant comparisons.

#### S11. H4 design, feasibility rules, and corrected-stage membership audit

H4 was post-hoc. The corrected-stage rerun reused the existing phenotype definitions, model forms, common covariates, survey design, medication-class composition definitions, profile registry, and multiplicity families. H4 eligibility assessments and representative-profile selection were completed without using new PTA4 outcome information and were fixed before new outcome access; this separation does not make H4 prospective or confirmatory because H4 itself was developed after earlier hearing results had been reviewed. Formal within-stage models required final stage-specific n ≥20, positive design degrees of freedom, the required categorical/reference levels, the expected phenotype-block degrees of freedom, a full-rank model matrix, finite coefficients and covariance matrix, and convergence. Meeting these requirements established computational estimability, not stable or precise inference.

**Table S17. Corrected-stage H4 participant membership audit**

| Participant | Historical stage | Corrected stage | H4-relevant eligibility | Effect on H4 membership |
|---|---|---|---|---|
| SEQN 119139 | 1 | 2 | Current waist observed; 10-year weight information missing. | Did not enter Stage 2 current-waist/weight-history model before or after correction. |
| SEQN 120761 | 0 | 1 | Current-waist/weight-history core complete. | Corrected Stage 1 remained outside H4 target stages. |

*Corrected-stage propagation did not change the retained H4 model-domain counts or final analytic sample sizes. The five multidimensional H4 test statistics and p values were unchanged.*

#### S12. H4 model specifications and survey support

The hypertension multidimensional block had four numerator df: reported hypertension duration, current SBP, medication-class composition (1 versus 2+ classes), and current-treatment duration. Medication-class composition entered the model as an observed phenotype marker and is not a measure of treatment intensity, hypertension severity, or resistant hypertension. Current-treatment duration was the within-person median across current relevant prescription records using RXDDAYS-derived duration information (National Center for Health Statistics 2021); it is a cross-sectional medication-history summary, not medication adherence, lifetime cumulative treatment, or a causal treatment effect. The current-waist/weight-history block had three numerator df: current waist circumference, measured current weight minus self-reported weight approximately 10 years earlier, and self-reported weight at age 25. H4 Wald tests used the model-domain design df as the denominator df. Variance handling followed the same Taylor-linearized survey design and lonely-PSU settings described above.

**Table S18. Survey-design support for the five multidimensional H4 models**

| Phenotype / stage | n | Strata | PSUs | Design df | Block df | Lonely strata | Model columns* |
|---|---|---|---|---|---|---|---|
| Hypertension / Stage 3 | 28 | 17 | 23 | 6 | 4 | 12 | 19 |
| Hypertension / Stage 4 | 29 | 18 | 24 | 6 | 4 | 12 | 20 |
| Waist/weight history / Stage 2 | 101 | 23 | 40 | 17 | 3 | 7 | 19 |
| Waist/weight history / Stage 3 | 58 | 21 | 32 | 11 | 3 | 11 | 19 |
| Waist/weight history / Stage 4 | 88 | 24 | 43 | 19 | 3 | 5 | 20 |

*Model columns include the intercept, phenotype block, and expanded common-covariate indicators. Each hypertension model contained 12 strata with only one PSU represented in the final domain. Full rank, convergence, and finite covariance confirm that the models are calculable. The representative-profile contrast CIs span −21.62 to 17.13 dB HL in Stage 3 and −13.07 to 28.07 dB HL in Stage 4, indicating substantial uncertainty. The model p values remain reported with this limitation.

#### S13. H4 parameter-level exploratory family

The six parameter-level tests formed a separate exploratory Holm multiplicity family. They were joint tests of stage-specific coefficients for one parameter while retaining the paired parameter in the same model. The hypertension model simultaneously included hypertension duration and current SBP; the diabetes model simultaneously included diabetes duration and HbA1c; the adiposity-history model simultaneously included current waist and derived 10-year weight change.

**Table S19. H4 parameter-level joint tests and conditional model structure**

| Parameter-level test | Joint model | n | df | F | Raw p | Holm-6 p |
|---|---|---|---|---|---|---|
| Hypertension duration | HTN duration + current SBP | 164 | 3, 23 | 0.777 | 0.519 | 1.000 |
| Current SBP | Current SBP + HTN duration | 164 | 3, 23 | 0.393 | 0.759 | 1.000 |

| Parameter-level test | Joint model | n | df | F | Raw p | Holm-6 p |
|---|---|---|---|---|---|---|
| Diabetes duration | Diabetes duration + HbA1c | 75 | 2, 16 | 5.109 | 0.0192 | 0.0962 |
| HbA1c | HbA1c + diabetes duration | 75 | 2, 16 | 2.105 | 0.154 | 0.617 |
| Waist circumference | Waist + derived 10-y weight change | 256 | 3, 25 | 22.295 | $3.05 \times 10^{-7}$ | $1.83 \times 10^{-6}$ |
| Derived 10-y weight change | 10-y weight change + waist | 256 | 3, 25 | 0.435 | 0.730 | 1.000 |

**Table S20. H4 parameter-level stage-specific slopes**

| Parameter | Stage | n | Slope, dB HL | 95% CI |
|---|---|---|---|---|
| HTN duration, +10 y | 2 | 52 | 0.923 | −3.04 to 4.89 |
| HTN duration, +10 y | 3 | 49 | −1.249 | −3.43 to 0.93 |
| HTN duration, +10 y | 4 | 63 | 2.051 | −2.35 to 6.45 |
| Current SBP, +10 mmHg | 2 | 52 | −1.390 | −4.38 to 1.60 |
| Current SBP, +10 mmHg | 3 | 49 | −0.211 | −2.72 to 2.30 |
| Current SBP, +10 mmHg | 4 | 63 | 0.278 | −2.27 to 2.82 |
| Diabetes duration, +10 y | 3 | 34 | −1.819 | −3.91 to 0.27 |
| Diabetes duration, +10 y | 4 | 41 | −3.513 | −6.81 to −0.21 |
| HbA1c, +1 point | 3 | 34 | −1.816 | −6.91 to 3.28 |
| HbA1c, +1 point | 4 | 41 | 2.313 | −0.18 to 4.81 |
| Waist, +10 cm | 2 | 102 | 5.093 | 3.42 to 6.77 |
| Waist, +10 cm | 3 | 63 | 0.906 | −1.40 to 3.21 |
| Waist, +10 cm | 4 | 91 | 2.601 | −1.05 to 6.25 |
| 10-y weight change, +5 kg | 2 | 102 | 0.018 | −1.12 to 1.15 |
| 10-y weight change, +5 kg | 3 | 63 | 0.559 | −0.81 to 1.93 |
| 10-y weight change, +5 kg | 4 | 91 | −0.651 | −2.64 to 1.34 |

The joint waist test evaluated whether all stage-specific waist slopes were zero while retaining derived 10-year weight change in the model. Its result does not establish an association with weight history. Self-reported weight at age 25 entered only the separate multidimensional current-waist/weight-history block in S14. Stage-specific slope CIs are descriptive; no individual slope p values were calculated.

#### S14. H4 multidimensional exploratory family

**Table S21. H4 multidimensional family-stage tests**

| Phenotype family / stage | n | Strata | PSUs | df | F | Raw p | Holm-5 p |
|---|---|---|---|---|---|---|---|
| Hypertension / Stage 3 | 28 | 17 | 23 | 4, 6 | 1.206 | 0.398 | 1.000 |
| Hypertension / Stage 4 | 29 | 18 | 24 | 4, 6 | 0.360 | 0.829 | 1.000 |
| Current waist/weight history / Stage 2 | 101 | 23 | 40 | 3, 17 | 9.759 | 0.000565 | 0.00283 |
| Current waist/weight history / Stage 3 | 58 | 21 | 32 | 3, 11 | 0.290 | 0.832 | 1.000 |
| Current waist/weight history / Stage 4 | 88 | 24 | 43 | 3, 19 | 2.980 | 0.0573 | 0.229 |

The Stage 2 current-waist/weight-history block met the five-test Holm criterion. This rejects the joint null for the three block coefficients. Because current waist and historical weight variables enter together, the test cannot attribute support specifically to weight history or establish Stage 2 specificity.

#### S15. Representative H4 profiles and descriptive contrasts

Representative profiles were exact observed phenotype vectors retained in the pre-existing H4 profile registry, which had been constructed from phenotype and support information and frozen before new outcome access within the H4 analysis; H4 itself remained post-hoc. For supported hypertension medication-class composition cells, the selected observed row minimized Euclidean distance from the componentwise median of the numeric phenotype features after scaling each feature by its sample standard deviation; R's first minimum resolved ties. For adiposity profiles, the target vectors were the 25th and 75th percentiles of the numeric phenotype features (R quantile type = 2), and the selected observed row minimized the same standard-deviation-scaled Euclidean distance, again using the first minimum for ties. Candidate cells required n ≥8 and nominal design df ≥1. Predictions fixed phenotype features to the selected observed vector, retained the final model-domain covariate distribution and survey weights, averaged the corresponding model-matrix rows, and propagated the survey::svyglm covariance matrix by the delta method. Reported intervals are pointwise survey-model intervals; no profile p values were calculated. The retained registry, rather than hearing outcomes, determined the reported profiles.

**Table S22. H4 hypertension representative profiles**

| Profile | Stage | HTN duration, y | SBP, mmHg | Medication-class composition | Current-treatment duration, y | Adjusted PTA4 (95% CI) |
|---|---|---|---|---|---|---|
| A | 3 | 18 | 138.0 | 1 antihypertensive class | 2.00 | 31.90 (20.07 to 43.73) |
| B | 3 | 28 | 145.0 | 2+ antihypertensive classes | 3.00 | 29.66 (12.03 to 47.29) |

| Profile | Stage | HTN duration, y | SBP, mmHg | Medication-class composition | Current-treatment duration, y | Adjusted PTA4 (95% CI) |
|---|---|---|---|---|---|---|
| A | 4 | 11 | 136.3 | 1 antihypertensive class | 10.99 | 31.79 (21.15 to 42.43) |
| B | 4 | 32 | 131.0 | 2+ antihypertensive classes | 9.99 | 39.29 (27.27 to 51.31) |

Profile A and Profile B label the first and second displayed profiles within each stage, respectively; these are reporting labels and do not define analytic factor levels. Within each stage, the two hypertension profiles differ simultaneously across all four displayed dimensions. The Stage 3 contrast was −2.24 dB HL (95% CI, −21.62 to 17.13) and the Stage 4 contrast was +7.50 dB HL (95% CI, −13.07 to 28.07). The wide intervals indicate substantial uncertainty in the profile contrasts. Because each pair differs simultaneously in four phenotype features, neither contrast identifies the association of an individual feature with PTA4.

**Table S23. H4 waist/weight-history representative profiles**

| Stage | Profile | Waist, cm | 10-y weight change, kg | Weight at age 25, kg | Adjusted PTA4 (95% CI) |
|---|---|---|---|---|---|
| 2 | Lower | 89.8 | −5.45 | 55.79 | 24.26 (20.29 to 28.22) |
| 2 | Higher | 106.4 | 2.98 | 70.31 | 31.96 (28.27 to 35.64) |
| 3 | Lower | 92.4 | −6.86 | 58.97 | 28.64 (21.16 to 36.12) |
| 3 | Higher | 111.2 | 1.86 | 72.57 | 31.90 (27.12 to 36.69) |
| 4 | Lower | 96.5 | −4.01 | 58.06 | 31.17 (27.20 to 35.14) |
| 4 | Higher | 115.8 | 5.79 | 79.38 | 33.88 (28.49 to 39.27) |

*The weight-history measures are self-reported or derived and should not be interpreted as measured longitudinal weight trajectories.*

**Table S24. H4 descriptive profile contrasts**

| Family | Stage | Contrast | Difference, dB HL | 95% CI |
|---|---|---|---|---|
| Hypertension | 3 | Profile B minus Profile A (Table S22) | −2.24 | −21.62 to 17.13 |
| Hypertension | 4 | Profile B minus Profile A (Table S22) | +7.50 | −13.07 to 28.07 |
| Current waist/weight history | 2 | Higher minus lower | +7.70 | 3.03 to 12.37 |
| Current waist/weight history | 3 | Higher minus lower | +3.26 | −4.49 to 11.01 |
| Current waist/weight history | 4 | Higher minus lower | +2.71 | −3.36 to 8.78 |

The Stage 2 7.70-dB contrast is a joint difference between two observed phenotype vectors. Its interpretation remains joint because the profiles differ in current waist and weight-history measures. The Stage 4 hypertension contrast likewise reflects simultaneous differences in reported duration, current SBP, medication-class composition, and current-treatment duration.

#### S16. H4 treatment-phenotype feasibility

Prescription products were decomposed at the ingredient level using official Multum ingredient categories. Verified hypertension classes were ACE inhibitor, angiotensin receptor blocker, beta blocker, calcium channel blocker, thiazide/thiazide-like diuretic, loop diuretic, potassium-sparing/mineralocorticoid antagonist, direct renin inhibitor, central sympatholytic, alpha blocker, and direct vasodilator. Broad or unspecified antihypertensive categories were treated as ambiguous and did not establish a unique class; categories outside the predefined medication-class dictionary were excluded from class counts. For the candidate three-level medication-class grouping, each stage-specific grouping cell was required to contain at least 8 participants and nominal design df ≥1; the same minimum thresholds were used for candidate representative-profile cells. The three-or-more-class antihypertensive cells had n = 1, 5, and 2 in Stages 2, 3, and 4, respectively, and therefore failed the retained cell-support criterion. The retained hypertension models used one class versus two-or-more classes. This treatment variable is an observed phenotype marker rather than a definition of resistant hypertension, treatment intensity, or hypertension severity.

**Table S25. Hypertension medication-class composition support audit**

| Treatment pattern | Stage 2 n | Stage 3 n | Stage 4 n | Support rule passed in all stages? |
|---|---|---|---|---|
| One antihypertensive class | 18 | 18 | 19 | Yes |
| Two antihypertensive classes | 8 | 9 | 10 | Yes |
| Three-or-more classes | 1 | 5 | 2 | No |

Because the three-or-more-class cells failed the n ≥8, nominal-df ≥1 cell-support criterion, a formal three-level medication-class composition hearing model was not fitted. The RXDDAYS-derived treatment-duration summary describes current prescription duration and contains no measure of adherence, cumulative exposure, or maximally tolerated dosing.

The diabetes treatment audit evaluated progressively simplified models incorporating treatment variables, insulin use, diabetes duration, glycemic burden, and treatment duration. No such diabetes hearing model met the sample-support and model-feasibility requirements for the post-hoc H4 analysis. This documents non-estimability and does not quantify a treatment-phenotype association.

#### S17. Secondary H4 kidney and lipid tests

**Table S26. Secondary H4 kidney and lipid tests**

| Family | Stage | Tested phenotype structure | n | df | F | Raw p |
|---|---|---|---|---|---|---|
| Kidney phenotype | 4 | Kidney pattern | 92 | 3, 18 | 4.272 | 0.01918 |
| Lipid | 2 | Current lipid-lowering therapy (none vs any classifiable therapy) | 63 | 1, 14 | 1.769 | 0.2048 |
| Lipid | 3 | Lipid pattern | 65 | 3, 12 | 0.0911 | 0.9636 |
| Lipid | 4 | Current lipid-lowering therapy (none vs any classifiable therapy) | 70 | 1, 15 | 0.675 | 0.4243 |

These tests were outside the two main H4 Holm multiplicity families and retain raw p values only. Kidney pattern combined eGFR <60 mL/min/1.73 $m^2$ and UACR ≥30 mg/g into four categories: neither abnormality, albuminuria alone, reduced filtration alone, or both abnormalities. The Stage 3 lipid pattern combined triglycerides ≥150 mg/dL and low HDL-C (<40 mg/dL in men or <50 mg/dL in women) into four categories: neither abnormality, high triglycerides alone, low HDL-C alone, or both abnormalities. Stages 2 and 4 instead compared no current lipid-lowering therapy with any classifiable lipid-lowering therapy, using prescription-medication records. Table S26 therefore does not represent a common lipid exposure across stages. The nominal Stage 4 kidney result remains Supplement-only and does not support a main conclusion.

#### S18. Reproducibility, code access, and software

Analyses were conducted in R 4.6.1 (R Core Team 2026). Complex-survey analyses used survey version 4.5 (Lumley 2026), and PREVENT risk estimation used AHAprevent version 1.0.0 with the base total-CVD implementation (Khan et al. 2024; American Heart Association 2026). The analysis archive contains versioned code for CKM stage assignment, analytic eligibility, survey-design specifications, H1-H4 models, representative-profile selection, and reported diagnostic and robustness analyses. Retained analysis objects support checks of H1 eligibility and model integrity. Historical and corrected H1 results are retained for comparison of adjusted means, omnibus tests, and coefficient-block diagnostics; the reported results use the corrected outputs. A study-planning record is available on the Open Science Framework (doi:10.17605/OSF.IO/BCNV7). No permanent public DOI for the complete versioned code archive had been assigned when this Supplement was prepared. The corresponding author can supply the versioned analysis code and audit materials upon reasonable request.
